# Trends in SARS-CoV-2 seroprevalence among pregnant women attending first antenatal care visits in Zambia: a repeated cross-sectional survey, 2021-2022

**DOI:** 10.1101/2024.01.02.24300729

**Authors:** Elizabeth Heilmann, Tannia Tembo, Sombo Fwoloshi, Bupe Kabamba, Felix Chilambe, Kalubi Kalenga, Mpanji Siwingwa, Conceptor Mulube, Victoria Seffren, Carolyn Bolton-Moore, John Simwanza, Samuel Yingst, Ruchi Yadav, Eric Rogier, Andrew F. Auld, Simon Agolory, Muzala Kapina, Julie R. Gutman, Theodora Savory, Chabu Kangale, Lloyd B. Mulenga, Izukanji Sikazwe, Jonas Z. Hines

**Affiliations:** Public Health Institute, Oakland, CA, USA; U.S. Centers for Disease Control and Prevention, Lusaka, Zambia; Centre for Infectious Disease Research in Zambia, Lusaka, Zambia; Ministry of Health, Lusaka, Zambia; PATH, Lusaka, Zambia; University Teaching Hospital, Lusaka, Zambia; U.S. Centers for Disease Control and Prevention, Atlanta, GA, USA; Zambia National Public Health Institute, Lusaka, Zambia

**Keywords:** COVID-19/epidemiology, COVID-19 Serological Testing, SARS-CoV-2, Seroepidemiologic Studies, Public Health Surveillance, Epidemiology/trends, Pregnant Women, Zambia, Africa

## Abstract

**Background:** SARS-CoV-2 serosurveys help estimate the extent of transmission and guide allocation of COVID-19 vaccines. We measured SARS-CoV-2 seroprevalence among women attending ANC clinics to assess exposure trends over time in Zambia.

**Methods:** We conducted repeated cross-sectional surveys among pregnant women aged 15-49 years attending their first ANC visits in four districts of Zambia (two urban and two rural) during September 2021-September 2022. Serologic testing was done using a multiplex bead assay which detects IgG antibodies to the nucleocapsid protein and the spike protein receptor-binding domain (RBD). We calculated monthly SARS-CoV-2 seroprevalence by district. We also categorized seropositive results as infection alone, infection and vaccination, or vaccination alone based on COVID-19 vaccination status and anti-RBD and anti-nucleocapsid test results.

**Findings:** Among 8,304 participants, 5,296 (63.8%) were cumulatively seropositive for SARS-CoV-2 antibodies. SARS-CoV-2 seroprevalence primarily increased from September 2021 to September 2022 in three districts (Lusaka: 61.8-100.0%, Chongwe: 39.6-94.7%, Chipata: 56.5-95.0%), but in Chadiza, seroprevalence increased from 27.8% in September 2021 to 77.2% in April 2022 before gradually dropping to 56.6% in July 2022. Among 5,906 participants with a valid COVID-19 vaccination status, infection alone accounted for antibody responses in 77.7% (4,590) of participants.

**Interpretation:** Most women attending ANC had evidence of prior SARS-CoV-2 infection and most SARS-CoV-2 seropositivity was infection-induced. Capturing COVID-19 vaccination status and using a multiplex bead assay with anti-nucleocapsid and anti-RBD targets facilitated distinguishing infection-induced versus vaccine-induced antibody responses during a period of increasing COVID-19 vaccine coverage in Zambia. Declining seroprevalence in Chadiza may indicate waning antibodies and a need for booster vaccines. ANC clinics have a potential role in ongoing SARS-CoV-2 serosurveillance and can continue to provide insights into SARS-CoV-2 antibody dynamics to inform near real-time public health responses.

## Introduction

SARS-CoV-2 was first detected in Zambia on March 18, 2020.^1^ Since then, the Zambia National Public Health Institute (ZNPHI) has reported nearly 350,000 COVID-19 cases over four main epidemic waves.^2^ Confirmed case counts underreport the true extent of SARS-CoV-2 infections due to the high proportion of subclinical infections, limited testing supplies, and surveillance gaps. SARS-CoV-2 seroprevalence studies can help bridge the gap in understanding SARS-CoV-2 transmission. In Zambia, a survey of six districts found a pooled prevalence of 10.6% in July 2020 and estimated that 92 infections occurred for every reported case.^3^ Another study conducted in a peri-urban district in February 2021 measured a seroprevalence of 33.7%.^4^ No studies have been published measuring SARS-CoV-2 seroprevalence in Zambia following the Delta and Omicron waves.

Repeated cross-sectional surveys leveraging existing healthcare platforms can help monitor trends in seroprevalence during ongoing transmission while minimizing implementation costs associated with large-scale household surveys. Antenatal care (ANC) clinics have been utilized in several countries in Africa to monitor SARS-CoV-2 seroprevalence in a healthy population of women accessing healthcare services.^5–9^ In Ethiopia, SARS-CoV-2 surveillance conducted in ANC clinics between April 2020 and March 2021 first detected SARS-CoV-2 antibodies in June 2020 and peaked at 11.8% in February 2021.^10^ Repeated cross-sectional serosurveys in ANC clinics in Kenya found that most women were seropositive for SARS-CoV-2 antibodies by October 2021.^11^

Early SARS-CoV-2 serosurveys measured antibodies to estimate total infections, but COVID-19 vaccine rollout now clouds seroprevalence interpretations. Detectable antibodies may be the result of SARS-CoV-2 infection, COVID-19 vaccination, or both. Serologic assays with nucleocapsid and spike targets in combination with COVID-19 vaccination history can distinguish these groups in countries, like Zambia, where most vaccines administered target the spike protein.^12^ COVID-19 vaccines first became publicly available in Zambia in April 2021.^13^ After a slow rollout initially, the Ministry of Health announced achieving 70% coverage of the targeted population on November 1, 2022.^14^ Most COVID-19 vaccine doses received and distributed in Zambia have been the spike-targeting Janssen (61%) and Pfizer-BioNTech (20%) vaccines.^13^

In Zambia, 97% of women attend at least one ANC visit during pregnancy, and antenatal attendees have long served as a sentinel population for HIV, syphilis, and malaria surveillance.^15–17^ Whether this platform can be leveraged to monitor COVID-19 remains unknown. We aimed to estimate monthly seroprevalence of SARS-CoV-2 antibodies among pregnant women attending first antenatal care visits at selected health facilities in four districts of Zambia from September 2021 to September 2022.

## Methods

### Study setting and population

We conducted repeated cross-sectional surveys of women attending first ANC visits in four districts (Chadiza, Chipata, Chongwe, and Lusaka) of Zambia from September 2021 to September 2022. Data collection began after Zambia’s third epidemic wave caused by the Delta variant (May-August 2021) and encompassed the fourth COVID-19 wave driven by the Omicron variant (December 2021-March 2022). The four districts were purposefully selected to capture a range of urban (Chipata and Lusaka) and rural (Chadiza and Chongwe) communities and districts with points-of-entry into Zambia (e.g., international airport in Lusaka and land borders with Malawi and Mozambique in Chipata and Chadiza, respectively).

A sample size of 200 women per district per month was selected based on available resources and an acceptable margin of error for the expected range of SARS-CoV-2 seroprevalence estimates over the study period. Ten facilities per district were randomly selected using probability proportional to the average number of ANC enrollments per month among health facilities reporting an average of at least 19 antenatal enrollments per month in 2020. Each facility in Chipata, Chongwe, and Lusaka districts enrolled up to 20 participants per month among pregnant women aged 15-49 years attending their first ANC visits. A health worker provided information on the study during group counseling sessions when the health facility provided routine first-time ANC services. Pregnant women willing to participate were screened for eligibility and, if consenting, were enrolled in the study until 20 participants were recruited per facility each month. In Chadiza District, all women attending the first ANC were approached for participation as part of a larger malaria surveillance pilot project, and up to 200 samples were selected post-enrollment for SARS-CoV-2 serologic testing per month.^18^ (For months with >200 participants in Chadiza, within each month, records were ordered by facility and interview date and the first 20 participants’ specimens were tested for SARS-CoV-2. If a facility had fewer than 20 participants in a month, specimens from other sites in Chadiza were selected and tested based on interview date until the sample size of 200 was reached.) SARS-CoV-2 surveillance in Chadiza goes until July 2022 (instead of September 2022) when the malaria surveillance project ended. At all sites, women were screened for acute COVID-19 prior to enrollment, and those exhibiting symptoms were referred for COVID-19 testing and case management per government protocols and excluded from the study.

### Data collection and sample processing

Participants were administered an electronic questionnaire on tablets using the Open Data Kit platform (Get ODK Inc., San Diego, CA, USA) to gather demographic data, SARS-CoV-2 exposures, COVID-19 vaccination, and routine ANC test results (i.e., HIV, syphilis, hepatitis, and malaria). Possible SARS-CoV-2 exposures included participants or households prior positive SARS-CoV-2 test and sustained close contact with a suspected or known COVID-19 case. COVID-19 vaccination and prior COVID-19 statuses were self-reported. Study staff attempted to verify COVID-19 vaccination information from vaccination cards if participants had them available on the day of their ANC visit. From February 2022 onward, participants self-reported inter-district and international travel, frequency of visits to churches, funerals or weddings, markets, and indoor dining, and use of public transportation and face masks to provide a general sense of COVID-19 risk behaviors.

Whole blood was collected on filter paper as dried blood spots (DBS) using the same finger prick or venipuncture from routine ANC testing. HIV, syphilis, hepatitis B, and malaria tests were conducted according to national guidelines, subject to the availability of test kits. Chipata, Chongwe, and Lusaka district specimens were transported to a central laboratory for testing at University Teaching Hospital (Lusaka, Zambia) and Chadiza District specimens were transported to PATH laboratory (Lusaka, Zambia) for testing. Serologic testing was done using the FlexImmArray SARS-CoV-2 Human IgG Antibody Test (Tetracore Inc, Rockville, MD, USA) on the MAGPIX platform (Luminex Corp, Austin, TX, USA) according to manufacturer instructions with DBS preparation as described by Tartof et al.^19–20^ The assay was verified in-country in conjunction with the US CDC prior to its use. This multiplex bead assay has three SARS-CoV-2 targets: nucleocapsid (N) protein, the receptor binding domain (RBD) of the spike-1 protein, and a N-RBD fusion protein. Signal ratios were calculated for each target as the median fluorescent intensity of the protein divided by the average calibrator median fluorescent intensity from each plate. Samples with signal ratios ≥1.2 for all three targets were considered SARS-CoV-2 IgG positive with past SARS-CoV-2 exposure, and samples with a signal ratio ≤0.9 for any target were considered SARS-CoV-2 IgG negative. Signal ratios >0.9 and <1.2 were considered indeterminate and the samples were re-run. Valid results of the second test were retained in the final dataset. Samples with indeterminate results after re-testing were categorized as SARS-CoV-2 IgG negative. Samples with persistent quality control issues were removed from the final dataset.

### Statistical analysis

SARS-CoV-2 seroprevalence was estimated as the number of positive cases divided by the total number of tests with valid results by month and district. Seroprevalence was adjusted based on the assay sensitivity (89.8%) and specificity (100.0%) from an independent test.^21^ Trend analysis was conducted to compare SARS-CoV-2 seroprevalence with district-level COVID-19 case reports provided by ZNPHI.^2^ District-level populations exposed to SARS-CoV-2 were calculated using 2022 Census population projection estimates from the Zambia Statistics Agency and peak SARS-CoV-2 seroprevalence estimates during data collection.^22^ These peak estimates of persons exposed to SARS-CoV-2 were compared to the total number of reported COVID-19 cases at the end of the study to estimate the ratio of reported cases to total SARS-CoV-2 infections in each district.

In a sub-analysis, target-specific assay results and COVID-19 vaccination status were analyzed to group antibody responses into no evident response (anti-RBD negative), infection only (anti-RBD positive, unvaccinated), vaccination only (anti-RBD positive, vaccinated, anti-N negative), or vaccination and infection (anti-RBD positive, vaccinated, anti-N positive) based off the decision tree developed by Duarte et al.^12^ Antibody responses in participants who received an unknown or inactivated virus vaccine were classified as indeterminate. Wilcoxon rank sum tests were used to compare signal ratios for each assay target by antibody response category. Logistic regression controlling for enrollment month was used to identify factors associated with seropositivity and calculate adjusted odds ratios. Sub-analyses were conducted among participants who reported prior confirmed COVID-19 and those who had received a COVID-19 vaccine. Analyses were conducted using R version 4.2.1 (R Foundation for Statistical Computing, Vienna, Austria).

## Results

Between September 2021 and September 2022, 9,221 pregnant women attending their first ANC visits were enrolled. A total of 8,558 (92.8%) samples were tested; among those, 8,304 (97.0%) valid SARS-CoV-2 antibody results were matched to participant questionnaires (Chadiza: 1,616, Chipata: 2,099, Chongwe: 2,441, Lusaka: 2,148). Two-hundred and fifty four specimens could not be linked to questionnaire data, so were not included in the analysis.

The median age of participants was 25 years (interquartile range [IQR]: 20-30; supplementary table 1). The majority (57.9%) of participants achieved primary education or less and most were unemployed (40.2%) or farmers (24.9%). Only 2.2% of women reported having COVID-19 prior to study enrollment, 3.3% reported confirmed COVID-19 in the household, and 3.9% reported close contact with a confirmed or suspected COVID-19 case outside the household. About half of participants responded that they wore a mask all or most of the time in public (49.5%) or were observed to be wearing a face mask properly during the ANC visit (49.3%). Vaccination coverage of one or more COVID-19 vaccine doses increased from 4.4% in September 2021 to 26.6% in September 2022. Most vaccinated women (1,274/1,688) had received the Janssen vaccine and 87.4% were considered fully vaccinated with the primary series.

Routine ANC testing for syphilis and hepatitis B was reported in less than half of participants (40.2% and 11.7%, respectively), but most (96.7%) women were tested for HIV or knew their HIV status prior to the visit and 8.8% were positive. Overall, 5,296 (63.8%) participants were SARS-CoV-2 seropositive. Seroprevalence was highest in Lusaka and lowest in Chadiza throughout the study period (figure 1). In Lusaka, SARS-CoV-2 adjusted seroprevalence rose from 61.8% (95% confidence interval [CI]: 52.8-70.5%) in September 2021 to 100.0% in August and September 2022 (August 95% CI: 97.2-100.0; September 95% CI: 100.0-100.0). SARS-CoV-2 seroprevalence also peaked near 100% in August 2022 in Chipata (97.3% [91.7-100.0]) and Chongwe (96.8% [91.9-100.0]) districts but dropped slightly in September 2022 (95.1% [89.0-99.7] and 94.7% [89.6-98.8], respectively). In Chadiza, SARS-CoV-2 seroprevalence peaked at 77.2% (95% CI: 61.8-78.6) in April 2022 and then gradually dropped to 56.6% (95% CI: 46.7-66.5) when data collection ended in July 2022. The greatest single month increase in urban districts (Lusaka and Chipata) was from December 2021 to January 2022 during the peak of the Omicron wave in Zambia, while in rural districts (Chadiza and Chongwe), the greatest increases were seen in March and April 2022, respectively.

**Figure 1.**
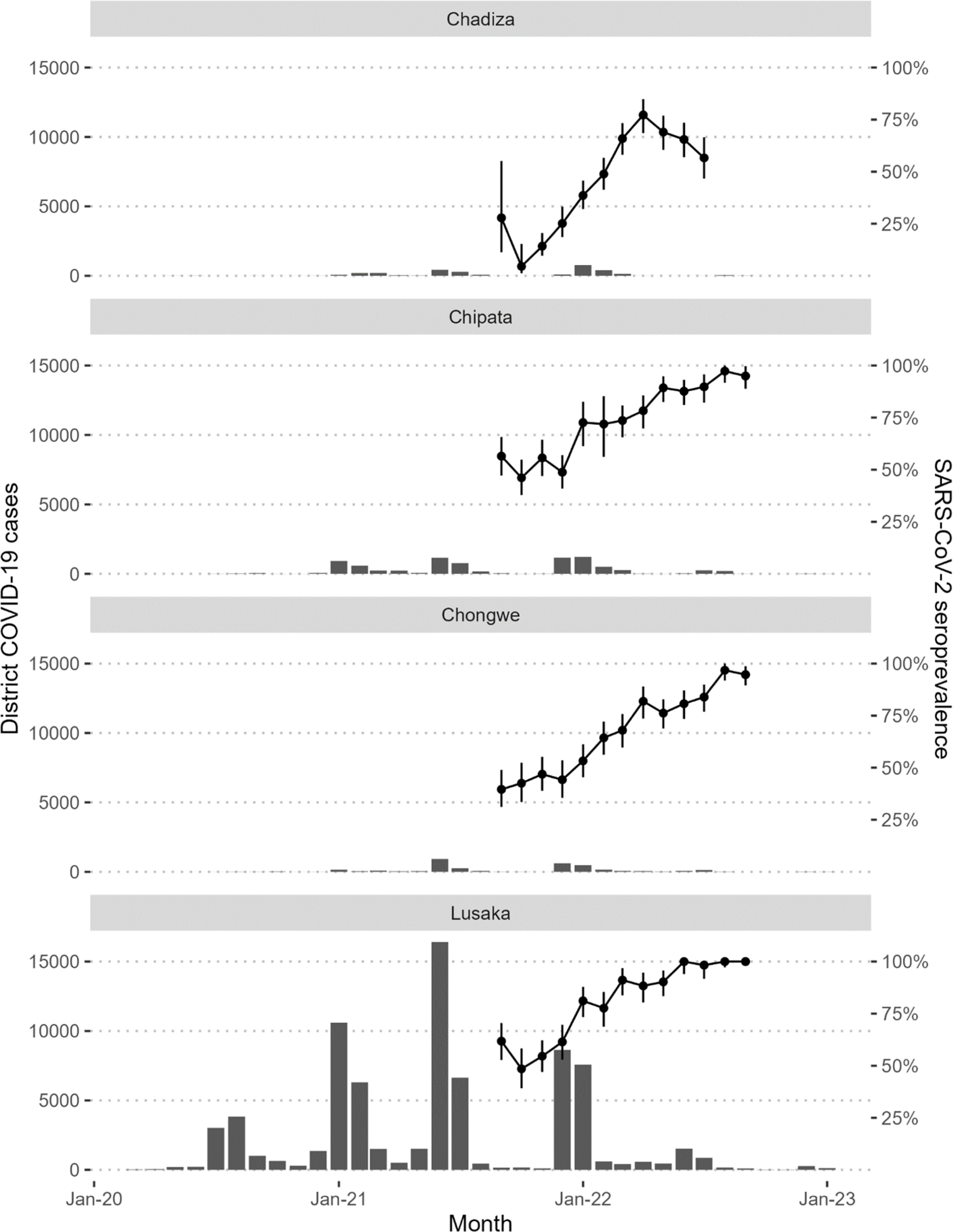
Reported COVID-19 cases among the general population and SARS-CoV-2 seroprevalence among study participants, by district. Serologic testing was done using the Tetracore FlexImmArray SARS-CoV-2 Human IgG Antibody Test; a positive result was assigned if signal ratios ≥1.2 for all 3 assay targets (nucleocapsid [N], receptor-binding domain [RBD] of the spike protein, and N-RBD fusion). Seroprevalence was adjusted for assay performance (sensitivity=89.8% and specificity=100%) and error bars represent 95% confidence intervals.

In the sub-analysis exploring target-specific assay results and COVID-19 vaccination status, 6,042 (72.8%) were positive for anti-RBD IgG and 6,343 (76.4%) were positive for anti-N IgG. Overall, 2,053 (24.7%) participants had no detectible antibodies to the RBD or nucleocapsid proteins, and 345 (4.2%) were indeterminate (supplementary figure 1). Among 5,906 participants with anti-RBD antibodies and a valid COVID-19 vaccination status, most (77.7%) were likely infection-induced only, 20.6% from vaccination and infection, and only 1.6% vaccination-induced alone. Infection-induced antibodies were the most common throughout the study period while hybrid infection and vaccination antibody responses became more common over time (figure 2). In Chadiza, hybrid antibodies surpassed infection-induced starting in April 2022. In comparing target-specific signal ratios by antibody response category, no difference was observed for the nucleocapsid target (figure 3). For the RBD target, the median signal ratio was significantly higher among participants who were vaccinated and infected compared to participants who were infected or vaccinated alone, and the median signal ratio for participants who were only infected was greater than for those who were only vaccinated.

**Figure 2.**
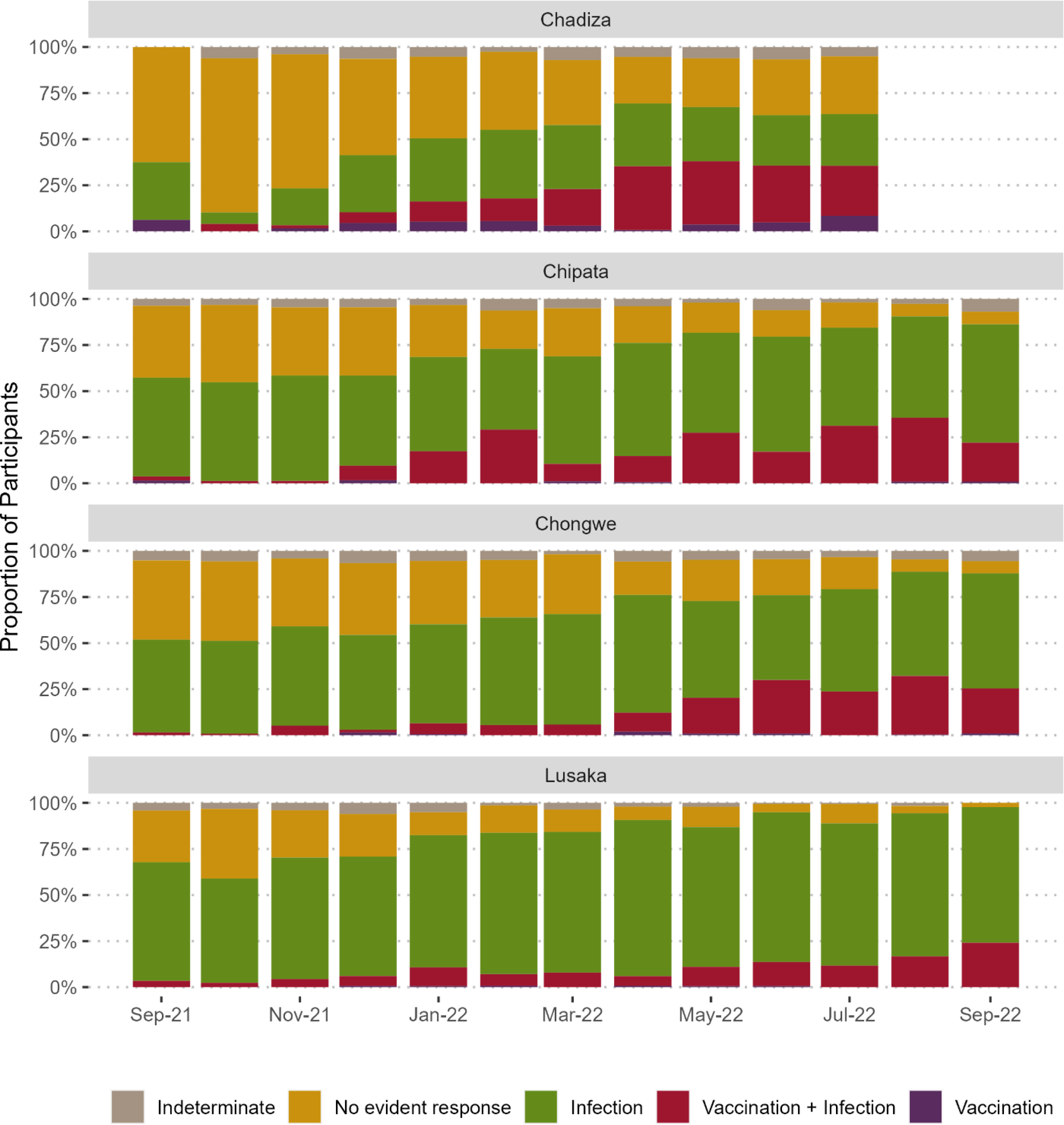
SARS-CoV-2 antibody response categorization of study participants by enrollment month and district. No evident antibody response was defined as testing negative for anti-RBD IgG. Infection-derived antibody response was defined as positive anti-RBD IgG AND no reported COVID-19 vaccination. Vaccination and infection-derived hybrid antibody response was defined as positive anti-RBD IgG AND positive anti-nucleocapsid IgG AND reported COVID-19 vaccination. Vaccination-derived antibody response was defined as positive anti-RBD IgG AND reported COVID-19 vaccination AND negative anti-nucleocapsid IgG. Equivocal IgG responses, unknown COVID-19 vaccination status, and unknown or Sinopharm vaccine type were categorized as indeterminate.

**Figure 3.**
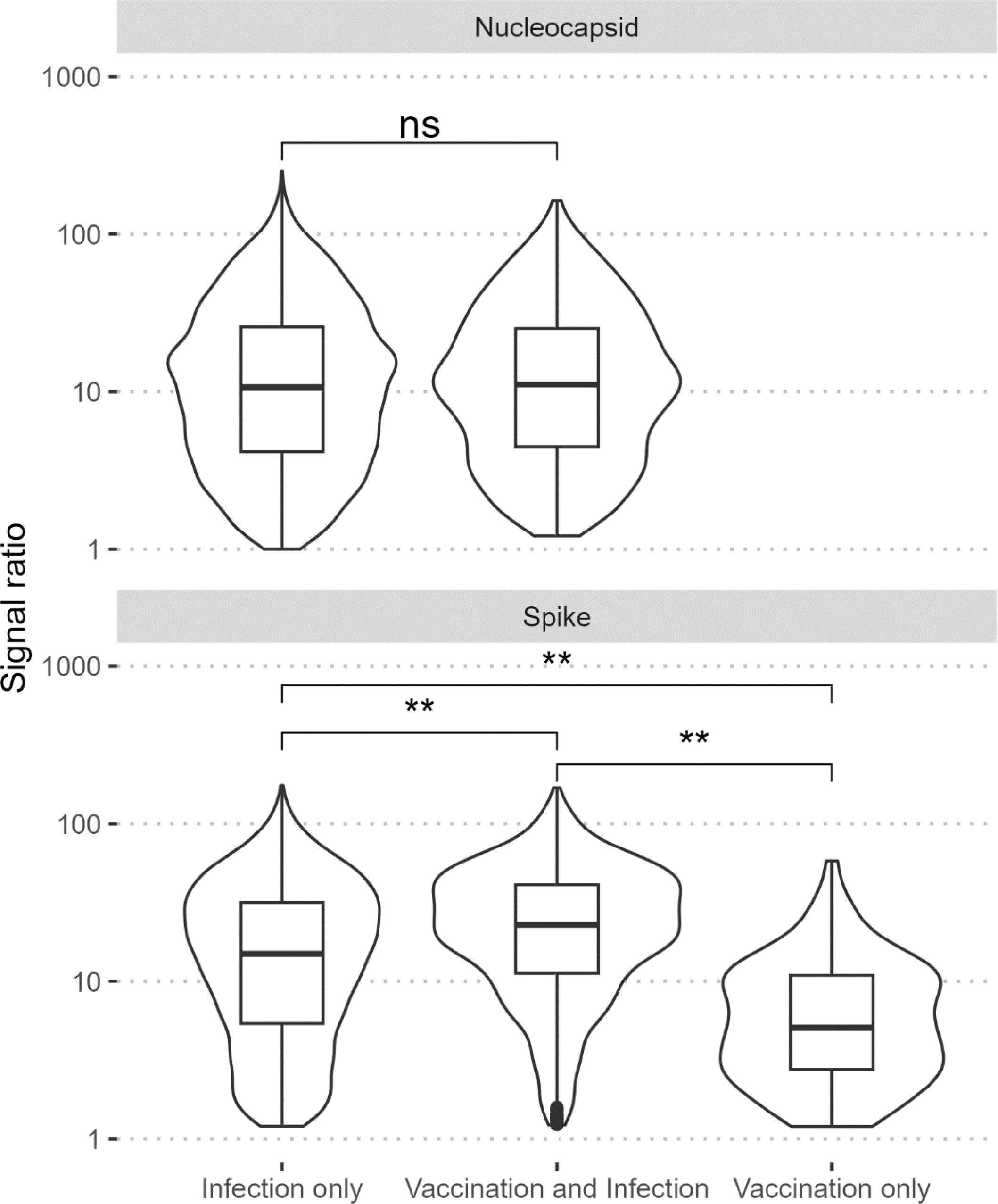
Signal ratios for three assay targets by antibody response category. Signal ratios were calculated by dividing the mean fluorescent intensity of the target by the mean fluorescent intensity of the calibrators. Vaccination and infection combined produced the strongest antibody response against all three assay targets compared to infection or vaccination alone. Participants whose antibody response was derived from infection only had greater signal ratios for the RBD target compared to participants with vaccination-derived antibody response only. ns=not significant (p-value>0.05); **p-value<0.001

Pairwise logistic regression adjusting for enrollment month identified different factors associated with SARS-CoV-2 serostatus in each district (table 1). Women aged 40-49 years in Chipata had nearly three times the odds of seropositivity compared to women aged 15-19 years (aOR: 2.76 [95% CI: 1.16-6.58]), but age was not significantly associated with serostatus elsewhere. In Chadiza and Chongwe districts, women who had completed secondary education or higher had greater odds of SARS-CoV-2 seropositivity compared to women with some primary education (Chadiza aOR: 1.76 [1.17-2.64]; Chongwe aOR: 1.64 [1.16-2.31]). Significant differences were observed in SARS-CoV-2 seroprevalence by occupation in all districts except Chadiza. In Chipata, women living with HIV (WLHIV) had higher odds of seropositivity compared to HIV-negative women (aOR: 1.42 [1.05-1.90]), although this association was no longer significant after further adjusting for age group (aOR: 1.29 [0.95-1.76]).

**Table 1.** SARS-CoV-2 seropositivity by participant demographics and SARS-CoV-2 exposures and risk behaviors.

|  | Chadiza |  | Chipata |  | Chongwe |  | Lusaka |  |
| --- | --- | --- | --- | --- | --- | --- | --- | --- |
|  | SARS-CoV-2 Seropositive | Adjusted Odds Ratio | SARS-CoV-2 Seropositive | Adjusted Odds Ratio | SARS-CoV-2 Seropositive | Adjusted Odds Ratio | SARS-CoV-2 Seropositive | Adjusted Odds Ratio |
|  | n (%) | aOR 95% CI | n (%) | aOR 95% CI | n (%) | aOR 95% CI | n (%) | aOR 95% CI |
| Total | 707 (43.8) |  | 1433 (68.3) |  | 1565 (64.1) |  | 1591 (74.1) |  |
| Age |  |  |  |  |  |  |  |  |
| 15-19 | 182 (41.6) | 1 (ref) | 244 (66.1) | 1 (ref) | 330 (63.7) | 1 (ref) | 154 (73.3) | 1 (ref) |
| 20-29 | 343 (45.1) | 1.23 (0.96-1.58) | 808 (67.9) | 1.12 (0.86-1.46) | 823 (63.3) | 1.00 (0.80-1.25) | 947 (73.0) | 0.98 (0.69-1.40) |
| 30-39 | 114 (45.4) | 1.12 (0.80-1.56) | 338 (70.3) | 1.31 (0.96-1.79) | 352 (66.3) | 1.21 (0.92-1.59) | 443 (76.9) | 1.22 (0.83-1.80) |
| 40-49 | 12 (46.2) | 1.43 (0.61-3.33) | 36 (83.7) | 2.76 (1.16-6.58)* | 60 (66.7) | 1.17 (0.70-1.95) | 42 (73.7) | 0.98 (0.48-2.00) |
| Education |  |  |  |  |  |  |  |  |
| No education | 156 (41.4) | 0.91 (0.70-1.18) | 108 (63.2) | 0.86 (0.59-1.26) | 44 (57.1) | 1.17 (0.70-1.95) | 16 (51.6) | 0.44 (0.19-1.00) |
| Some primary | 337 (43.3) | 1 (ref) | 329 (63.5) | 1 (ref) | 485 (60.0) | 1 (ref) | 296 (71.7) | 1 (ref) |
| Completed primary | 49 (36.8) | 0.96 (0.64-1.43) | 390 (70.5) | 1.08 (0.82-1.41) | 405 (66.6) | 1.12 (0.89-1.42) | 255 (75.7) | 0.91 (0.64-1.30) |
| Some secondary | 95 (47.5) | 1.23 (0.88-1.70) | 440 (71.9) | 1.25 (0.96-1.63) | 485 (66.3) | 1.34 (1.07-1.67) | 602 (74.8) | 0.94 (0.70-1.25) |
| Completed secondary or higher | 68 (55.3) | 1.76 (1.17-2.64)* | 161 (67.1) | 1.14 (0.81-1.61) | 146 (67.9) | 1.64 (1.16-2.31)* | 420 (75.1) | 0.96 (0.71-1.31) |
| Occupation |  |  |  |  |  |  |  |  |
| Farmer | 596 (43.1) | 0.63 (0.37-1.10) | 274 (61.9) | 0.66 (0.51-0.85)* | 349 (58.0) | 0.70 (0.56-0.87)* | 9 (90.0) | 2.55 (0.30-21.85) |
| Daily labourer/pieceworker | 4 (50.0) | 1.20 (0.25-5.81) | 49 (62.8) | 0.76 (0.46-1.26) | 205 (63.1) | 0.88 (0.67-1.16) | 55 (85.9) | 1.93 (0.90-4.15) |
| Merchant/shop owner | 3 (25.0) | 0.25 (0.06-1.08) | 30 (53.6) | 0.55 (0.31-0.98)* | 33 (60.0) | 1.08 (0.6-1.95) | 83 (76.9) | 1.94 (1.19-3.18)* |
| Health worker | 4 (50.0) | 1.11 (0.24-5.10) | 8 (50.0) | 0.50 (0.17-1.42) | 5 (83.3) | 4.70 (0.48-45.97) | 10 (76.9) | 1.60 (0.40-6.40) |
| Government/civil servant | 13 (48.1) | 0.83 (0.32-2.19) | 35 (74.5) | 1.16 (0.58-2.33) | 15 (88.2) | 5.02 (1.07-23.47)* | 61 (69.3) | 0.91 (0.55-1.52) |
| Private sector employee | 3 (50.0) | 0.66 (0.11-3.95) | 21 (67.7) | 0.91 (0.41-2.02) | 20 (80.0) | 3.38 (1.19-9.58)* | 107 (73.8) | 1.11 (0.73-1.70) |
| Hawker/market seller | 6 (42.9) | 1.05 (0.30-3.67) | 102 (71.8) | 1.00 (0.67-1.51) | 121 (73.8) | 1.56 (1.05-2.34)* | 181 (75.1) | 1.20 (0.86-1.69) |
| Student | 42 (47.2) | 0.61 (0.30-1.23) | 70 (73.7) | 1.16 (0.70-1.91) | 54 (68.4) | 1.15 (0.69-1.94) | 88 (79.3) | 0.94 (0.56-1.58) |
| Unemployed | 34 (54.0) | 1 (ref) | 719 (72.2) | 1 (ref) | 712 (65.4) | 1 (ref) | 859 (71.9) | 1 (ref) |
| Other occupation | 0 (0.0) | -- -- | 102 (63.0) | 0.76 (0.53-1.10) | 37 (68.5) | 1.24 (0.66-2.33) | 129 (79.6) | 1.34 (0.87-2.06) |
| Prior COVID-19 |  |  |  |  |  |  |  |  |
| No | 678 (43.3) | 1 (ref) | 1389 (68.4) | 1 (ref) | 1525 (64.2) | 1 (ref) | 1518 (73.9) | 1 (ref) |
| Yes | 28 (66.7) | 3.06 (1.55-6.07)* | 28 (66.7) | 0.95 (0.48-1.89) | 22 (71.0) | 1.68 (0.72-2.95) | 52 (81.2) | 1.25 (0.64-2.45) |
| Household COVID-19 |  |  |  |  |  |  |  |  |
| No | 684 (43.5) | 1 (ref) | 1379 (68.5) | 1 (ref) | 1451 (63.8) | 1 (ref) | 1458 (73.5) | 1 (ref) |
| Yes | 17 (47.2) | 1.20 (0.60-2.38) | 31 (63.3) | 0.95 (0.51-1.77) | 36 (75.0) | 1.45 (0.71-2.95) | 112 (81.8) | 1.19 (0.75-1.91) |
| Close COVID-19 contact<sup>†</sup> |  |  |  |  |  |  |  |  |
| No | 629 (43.7) | 1 (ref) | 1348 (69.0) | 1 (ref) | 1277 (64.1) | 1 (ref) | 1365 (73.3) | 1 (ref) |
| Yes | 11 (36.7) | 0.93 (0.42-2.05) | 32 (61.5) | 0.80 (0.44-1.46) | 41 (66.1) | 0.95 (0.53-1.67) | 140 (79.5) | 1.10 (0.73-1.66) |
| COVID-19 vaccination<sup>†</sup> |  |  |  |  |  |  |  |  |
| No | 396 (36.4) | 1 (ref) | 1057 (64.7) | 1 (ref) | 1164 (60.5) | 1 (ref) | 1379 (72.4) | 1 (ref) |
| Yes | 299 (58.5) | 1.76 (1.40-2.21)* | 363 (80.3) | 1.48 (1.13-1.93)* | 381 (78.4) | 1.40 (1.09-1.80)* | 208 (88.1) | 1.97 (1.29-3.02)* |

| HIV status |  |  |  |  |  |  |  |  |
| --- | --- | --- | --- | --- | --- | --- | --- | --- |
| Negative | 767 (44.0) | 1 (ref) | 1212 (67.5) | 1 (ref) | 1404 (65.4) | 1 (ref) | 1388 (75.3) | 1 (ref) |
| Positive | 16 (33.3) | 0.83 (0.43-1.59) | 203 (72.8) | 1.42 (1.05-1.90)* | 113 (52.6) | 0.77 (0.57-1.05) | 104 (63.4) | 0.87 (0.60-1.26) |
| aOR = adjusted odds ratio (adjusted for enrollment month); CI = confidence interval. *Close contact defined as being within 2 meters of someone outside the household suspected or confirmed to have COVID-19 for greater than 15 minutes; †≥1 dose of any COVID-19 vaccine. *p-value<0.05 |  |  |  |  |  |  |  |  |

SARS-CoV-2 seropositivity was significantly associated with prior confirmed COVID-19 only in Chadiza (aOR=3.06 [1.55-6.07]; table 1). Among 179 reported prior COVID-19 cases, 130 (72.6%) were SARS-CoV-2 seropositive at the time of enrollment. The median time between COVID-19 diagnosis and first ANC visit was 9 months (IQR: 7-14) among women who tested SARS-CoV-2 seropositive and 8 months (IQR: 4-13) among seronegative women (p = 0.13).

COVID-19 vaccination was associated with significantly higher odds of SARS-CoV-2 seropositivity in all four districts, adjusting for enrollment month (table 1). Among vaccinated women, SARS-CoV-2 serostatus did not differ by partial vs. full primary vaccine series or vaccine type (table 2). Compared to women who received their first COVID-19 vaccine dose within one month prior to study enrollment, participants who received their first dose one to five months prior had greater odds of testing SARS-CoV-2 seropositive, controlling for enrollment month and district (aOR: 1.58 [1.19-2.10]).

**Table 2.** SARS-CoV-2 seroprevalence among vaccinated participants (n=1685)

|  |  |  |  |  |  |  |
| --- | --- | --- | --- | --- | --- | --- |
|  | SARS-CoV-2 Seropositive |  | SARS-CoV-2 Seronegative |  | Adjusted Odds Ratio |  |
|  | n | (%) | n | (%) | aOR | (95% CI) |
| Vaccination status |  |  |  |  |  |  |
| Partially vaccinated | 123 | (70.3) | 52 | (29.7) | 0.74 | (0.51-1.07) |
| Full primary series | 1106 | (74.9) | 370 | (25.1) | 1 | (ref) |
| Vaccine type |  |  |  |  |  |  |
| Oxford/AstraZeneca (ChAdOx1-S) | 150 | (64.9) | 81 | (35.1) | 1 | (ref) |
| J&J/Janssen (Ad.26.COV2.S) | 965 | (75.7) | 309 | (24.3) | 1.28 | (0.92-1.79) |
| Pfizer-BioNTech (BNT162b2) | 50 | (75.8) | 16 | (24.2) | 0.71 | (0.36-1.39) |
| Sinopharm | 32 | (72.7) | 12 | (27.3) | 0.79 | (0.37-1.69) |
| Mixed | 21 | (91.3) | 2 | (8.7) | 2.82 | (0.60-13.26) |
| Other | 7 | (100.0) | 0 | (0.0) | -- | -- |
| Months between first dose and enrollment |  |  |  |  |  |  |
| <1 month | 290 | (69.5) | 127 | (30.5) | 1 | (ref) |
| 1-5 months | 669 | (75.8) | 214 | (24.2) | 1.58 | (1.19-2.10)* |
| ≥6 months | 223 | (81.4) | 51 | (18.6) | 1.48 | (0.99-2.22) |
| aOR=adjusted odds ratio (adjusted for district and enrollment month); CI=confidence interval. *p<0.05 |  |  |  |  |  |  |

Twenty-one percent of the 179 participants with prior COVID-19 reported being hospitalized. Hospitalization and time between COVID-19 and enrollment were not associated with SARS-CoV-2 serostatus, but COVID-19 vaccination was associated with greater odds of seropositivity among these women (OR: 4.32 [2.07-9.02]; table 3).

**Table 3.** Characteristics participants reporting prior COVID-19 by SARS-CoV-2 serostatus (n=179)

|  |  |  |  |
| --- | --- | --- | --- |
|  | SARS-CoV-2<br>Seropositive | SARS-CoV-2<br>Seronegative | Adjusted Odds Ratio |
|  | n (%) | n (%) | aOR (95% CI) |
| Hospitalized due to COVID-19 |  |  |  |
| No | 100 (78.7) | 39 (79.6) | 1 (ref) |
| Yes | 27 (21.3) | 10 (20.4) | 0.63 (0.24-1.66) |
| Months between diagnosis and enrollment |  |  |  |
| <6 months | 20 (18.3) | 13 (31.0) | 1 (ref) |
| 6-11 months | 41 (37.6) | 12 (28.6) | 1.02 (0.37-2.79) |
| ≥12 months | 48 (44.0) | 17 (40.5) | 0.41 (0.11-1.51) |
| COVID-19 vaccination<sup>†</sup> |  |  |  |
| No | 54 (41.9) | 37 (75.5) | 1 (ref) |
| Yes | 75 (58.1) | 12 (24.5) | 4.11 (1.80-9.41)* |
| aOR=adjusted odds ratio (adjusted for district and enrollment month); CI=confidence interval. <sup>†</sup>Received ≥1 doses of any COVID-19 vaccine. *p<0.05 |  |  |  |

District-level cumulative incidence was calculated from 2022 population projections and reported COVID-19 cases from March 2020 through the end of the survey (July 2022 for Chadiza and September 2022 elsewhere) and ranged from 1.1% in Chadiza District to 3.5% in Lusaka District (table 4). Overall, 90,150 COVID-19 cases were reported in the four districts from March 2020 to September 2022 but we estimated that 2,911,393 people had evidence of SARS-CoV-2 exposure. The ratio of SARS-CoV-2 seroprevalence to cumulative incidence was 29:1 in Lusaka District, 31:1 in Chadiza District, 39:1 in Chipata District, and 92:1 in Chongwe District.

**Table 4.** Estimated populations exposed to SARS-CoV-2 and ratios of peak seroprevalence to cumulative incidence during last survey month.

|  |  |  |  |  |
| --- | --- | --- | --- | --- |
|  | Chadiza | Chipata | Chongwe | Lusaka |
| Peak SARS-CoV-2 seroprevalence<sup>†</sup> | 77.2% | 97.3% | 96.8% | 100.0% |
| 2022 census population | 111,069 | 327,059 | 313,389 | 2,204,059 |
| Reported COVID-19 cases | 2,761 | 8,096 | 3,303 | 75,990 |
| Cumulative incidence<sup>‡</sup> | 2.5% | 2.5% | 1.1% | 3.4% |
| Estimated population with evidence of SARS-CoV-2 exposure<sup>§</sup> | 85,745 | 318,228 | 303,361 | 2,204,059 |
| Ratio of seroprevalence to cumulative incidence | 31:1 | 39:1 | 92:1 | 29:1 |
| <sup>†</sup>Peak SARS-CoV-2 seroprevalence for Chadiza District was April 2022; for Chipata District, peak was August 2022; for Chongwe district, peak was August 2022; for Lusaka District, peak was September 2022. <sup>‡</sup>Calculated as reported cases divided by population projection. <sup>§</sup>Calculated as seroprevalence multiplied by population projection. 2022 census data were obtained from the Zambia Statistics Agency and reported COVID-19 cases were obtained from the Zambia National Public Health Institute. |  |  |  |  |

## Discussion

SARS-CoV-2 seroprevalence was high among participants and nearly all first ANC attendees in three out of four study districts had evidence of SARS-CoV-2 infection. In those districts, seroprevalence generally increased over the 13-month study period, while in Chadiza, it declined from April to July 2022. Lower seroprevalence in Chadiza District was expected, being the most rural of the study districts. The very high seroprevalence in Lusaka District was also expected given a study conducted at two Lusaka hospitals which found that 36.9% of women attending ANC clinics between March and July 2021 (part of the Delta wave, prior to this study) were positive for SARS-CoV-2 by RT-PCR.^23^ The large rise in SARS-CoV-2 seroprevalence from November 2021 to March 2022 when the Omicron wave occurred was similar to what was seen in South Africa where seroprevalence among the general population was 73% before the Omicron wave and 91% after.^24–25^ This high seroprevalence was proposed as a reason for the lack of significant transmission waves after Omicron, despite the emergence of Omicron sub-lineages with greater infectivity and low COVID-19 vaccine coverage.^25^

COVID-19 vaccination among participants was low and most SARS-CoV-2 antibodies were likely infection-induced. COVID-19 vaccination was associated with higher odds of SARS-CoV-2 seropositivity and COVID-19 vaccination in combination with SARS-CoV-2 infection produced the strongest immune response, both of which could be explained by a robust immune response caused by hybrid immunity.^26^ Other studies in Africa also measured higher SARS-CoV-2 seroprevalence among vaccinated participants, higher neutralizing antibody titers among participants with infection and vaccination, and faster waning of antibodies post-vaccination among participants who had not been infected.^7,27–29^ These results support the recommendation for COVID-19 vaccination (including booster doses) regardless of past infection. COVID-19 vaccination campaigns were conducted throughout Zambia during the study period, but vaccine coverage among study participants was lower than the general population, possibly reflecting hesitancy toward vaccination during pregnancy, and only about 12% of the fully vaccinated population has received a booster dose.^2^ COVID-19 messages and risk mitigation strategies addressing the concerns of pregnant women can be incorporated seamlessly into routine ANC to prevent adverse outcomes in this population at higher risk for severe COVID-19. Healthcare providers can screen for symptoms of COVID-19, malaria, and tuberculosis simultaneously, and COVID-19 vaccines may be administered alongside tetanus boosters.

The ratios of SARS-CoV-2 seroprevalence to cumulative incidence were lower than the 1:92 ratio previously measured in a July 2020 serosurvey in other parts of Zambia.^3^ This reduction mirrors the downward global trend in seroprevalence to cumulative incidence ratios since 2020 (i.e., improving case detection ratios) and may be due to evolving testing strategies and increased availability and use of rapid antigen test kits.^30^ Interpretation of these ratios late in the pandemic is complicated by reinfections, waning antibodies, and non-report of home RDT results, leading to an underestimation of total SARS-CoV-2 infections. Some past SARS-CoV-2 infections may not be detectable as antibodies wane over time and people with asymptomatic infections or mild clinical cases may mount less of an immune response.^31^ This is consistent with the women who were SARS-CoV-2 seronegative despite reporting prior COVID-19 and may explain the declining seroprevalence observed in some districts and months.

Older age, higher education, and urban versus rural location were all associated with higher seroprevalence during the July 2020 survey in Zambia, and similar associations were noted in some districts during this study.^3^ This may reflect differences in access to COVID-19 information and infection prevention supplies or in the ability to comply with COVID-19 risk mitigation guidelines among certain subpopulations. These factors may intersect with occupation which was also associated with differences in seroprevalence in most districts. Diverse work environments, such as open fields for farming, compact cubicles in an office, or crowded market stalls, could impact an individual’s SARS-CoV-2 exposure risk.

Similar associations were noted in Mozambique where market sellers and those engaged in formal employment had higher SARS-CoV-2 seroprevalence than other community members.^29^ Furthermore, modes of transportation might vary by urban/rural status, which could in turn affect risk of coming into contact with SARS-CoV-2. Tailored COVID-19 prevention messaging for demographic groups with higher SARS-CoV-2 seroprevalence may help reduce transmission during future waves. These observed differences in SARS-CoV-2 prevalence by certain demographic groups could simply reflect differences in COVID-19 risk for different groups in the population.

Most COVID-19 mitigation strategies in Zambia were carried out nationally by the Ministry of Health. Given the differences in SARS-CoV-2 risk factors and seroprevalence trends by district, subnational strategies may be more suitable in the future to mitigate COVID-19 risk where and when needed. District or provincial level responses could help direct resources to areas of high transmission while avoiding imposing strict measures in areas with low transmission where the social and economic burdens of mitigation efforts may outweigh the benefit of reducing transmission. This approach was used to lift masking requirements in Zambia as districts reached 70% vaccination coverage and demonstrates that robust surveillance systems and quality tracking of key COVID-19 indicators at the district level can facilitate subnational decision-making.^32^

This study has several limitations. First, we could not estimate a single population-weighted seroprevalence because of the non-systematic selection of districts and participants. Our study was limited to women aged 15-49 years and may not be representative of populations that may be more or less susceptible to SARS-CoV-2 infection. However, a global meta-regression analysis found no difference in seroprevalence by sampling frame comparing studies among pregnant women to household and community surveys.^30^ As with all SARS-CoV-2 seroprevalence surveys, our estimates depend on serologic test performance. The manufacturer’s requirement of testing positive for all three targets for an overall positive sample is more conservative than single target assays and may have excluded some past infections. Waning of SARS-CoV-2 antibodies might have resulted in underestimation of seroprevalence, and variability in the waning time for different antibody types could have biased the sub-analysis findings categorizing infection/vaccination statuses. Studies have shown that anti-nucleocapsid antibodies wane more quickly than anti-spike antibodies,^33–34^ although anti-N antibody prevalence was slightly greater than anti-RBD antibody prevalence in our study. Adjusting the seroprevalence estimates based on a test sensitivity of 90% likely helped but the potential for misclassification remains. Given the cross-sectional study design, we could not assess whether those who were seronegative had truly never been exposed to SARS-CoV-2, had been exposed and never mounted a detectable antibody response, or had sero-reverted before testing. Lastly, we were uncertain how the premature ending of SARS-CoV-2 surveillance in Chadiza in July 2022 affected the overall seroprevalence estimates for this study.

Despite these limitations, our study also has notable strengths. We measured SARS-CoV-2 seroprevalence over 13 months which encompassed periods before, during, and after Zambia’s Omicron wave. The study period also spanned early COVID-19 vaccine rollout in Zambia, and the use of a multiplex bead assay with spike and nucleocapsid targets allowed us to distinguish seropositivity from infection versus vaccination. We included several districts which had not been included in previous SARS-CoV-2 seroprevalence studies and found substantial geographic variation, particularly in the most rural study district. Finally, we demonstrated the feasibility of integrating SARS-CoV-2 serosurveillance into routine health services which may be adaptable to future epidemics.

As the COVID-19 pandemic continues to evolve, so does the role of seroprevalence studies. While our study shows that most people living in Zambia have likely been exposed to SARS-CoV-2, seroprevalence studies may still play a valuable role in understanding waning population immunity over time and susceptibility to emerging variants. Longitudinal cohort studies may be better suited than cross-sectional surveys to meet these needs, and ANC may continue to be a useful platform for recruiting participants as women attend frequent follow-up visits during pregnancy and postpartum periods.

## Author Contributions

EH, TT, SF, BK, CBM, SY, JRG, VS, LM, IS, and JZH designed the study. TT, BK, and KK monitored data and sample collection, transport, and storage. FC, MS, and CM performed the lab analysis, and VS, RY, and ER interpreted the lab results. EH, JS, MK, and JZH accessed and verified the underlying data. EH analyzed the data and produced the figures. EH wrote the first draft of the manuscript and all other coauthors reviewed and approved of the final version. All authors had access to the study data and approved the final copy of the manuscript for publication submission.

## Ethics statement

This study was authorized by the National Health Research Authority (NHRA) through an expedited review of COVID-19 related studies. For Chadiza District, the SARS-CoV-2 data collection and serologic testing were reviewed and approved by the University of Zambia Biomedical Research Ethics Committee as an amendment to a pre-existing malaria surveillance protocol. This activity was reviewed by CDC and was conducted consistent with applicable federal law and CDC policy (See e.g., 45 C.F.R. part 46.102(l)(2), 21 C.F.R. part 56; 42U.S.C. §241(d); 5 U.S.C. §552a;44 U.S.C. §3501 et seq). All methods were carried out in accordance with relevant guidelines and regulations. Written informed consent was obtained for all participants before enrollment. Pregnant women under 18 years are considered emancipated minors in Zambia for the purpose of providing their own informed consent.

## Data Availability Statement

The data that support the findings of this study are available from Zambia Ministry of Health, but restrictions apply to the availability of these data, which were used under license for the current study, and so are not publicly available. Data are however available from the authors upon reasonable request and with permission of Zambia Ministry of Health. Interested researchers should submit a research proposal to the CDC Zambia science office at. If approved, the requestor must sign a data use agreement. Additionally, the study protocol is available for request.

## Authorship Disclaimer

The findings and conclusions in this report are those of the author(s) and do not necessarily represent the official position of the U.S. Centers for Disease Control and Prevention.

---

**Supplementary table 1.** Participant demographics, SARS-CoV-2 exposures and risk behaviors, and ANC test results.

|  |  |  |  |  |  |
| --- | --- | --- | --- | --- | --- |
|  | Chadiza | Chipata | Chongwe | Lusaka | Total |
|  | n=1616 | n=2099 | n=2441 | n=2148 | N=8304 |
|  | n (%) | n (%) | n (%) | n (%) | n (%) |
| Age, median (IQR) | 22 (19-28) | 25 (21-30) | 24 (20-30) | 26 (22-30) | 25 (20-30) |
| 15-19 | 438 (27.1) | 369 (17.6) | 518 (21.2) | 210 (9.8) | 1535 (18.5) |
| 20-29 | 760 (47.0) | 1190 (56.7) | 1300 (53.3) | 1297 (60.4) | 4547 (54.8) |
| 30-39 | 251 (15.5) | 481 (22.9) | 531 (21.8) | 576 (26.8) | 1839 (22.1) |
| 40-49 | 26 (1.6) | 43 (2.0) | 90 (3.7) | 57 (2.7) | 216 (2.6) |
| Unknown | 141 (8.7) | 16 (0.8) | 2 (0.1) | 8 (0.4) | 167 (2.0) |
| Education |  |  |  |  |  |
| No education | 377 (23.3) | 171 (8.1) | 77 (3.2) | 31 (1.4) | 656 (7.9) |
| Some primary | 779 (48.2) | 518 (24.7) | 809 (33.1) | 413 (19.2) | 2519 (30.3) |
| Completed primary | 133 (8.2) | 553 (26.3) | 608 (24.9) | 337 (15.7) | 1631 (19.6) |
| Some secondary | 200 (12.4) | 612 (29.2) | 732 (30.0) | 805 (37.5) | 2349 (28.3) |
| Completed secondary or higher | 123 (7.6) | 240 (11.4) | 215 (8.8) | 559 (26.0) | 1137 (13.7) |
| Unknown | 4 (0.2) | 5 (0.2) | 0 (0.0) | 3 (0.1) | 12 (0.1) |
| Occupation |  |  |  |  |  |
| Farmer | 1383 (85.6) | 443 (21.1) | 602 (24.7) | 10 (0.5) | 2438 (24.9) |
| Daily laborer/pieceworker | 8 (0.5) | 78 (3.7) | 325 (13.3) | 64 (3.0) | 475 (5.7) |
| Merchant/shop owner | 12 (0.7) | 56 (2.7) | 55 (2.3) | 108 (5.0) | 231 (2.8) |
| Health worker | 8 (0.5) | 16 (0.8) | 6 (0.2) | 13 (0.6) | 43 (0.5) |
| Government employee/civil servant | 27 (1.7) | 47 (2.2) | 17 (0.7) | 88 (4.1) | 179 (2.2) |
| Private sector employee | 6 (0.4) | 31 (1.5) | 25 (1.0) | 145 (6.8) | 207 (2.5) |
| Hawker/market seller | 14 (0.9) | 142 (6.8) | 164 (6.7) | 241 (11.2) | 561 (6.8) |
| Student | 89 (5.5) | 95 (4.5) | 79 (3.2) | 111 (5.2) | 374 (4.5) |
| Unemployed | 63 (3.9) | 996 (47.5) | 1088 (44.6) | 1195 (55.6) | 3342 (40.2) |
| Other occupation | 1 (0.1) | 162 (7.7) | 54 (2.2) | 162 (7.5) | 379 (4.6) |
| Unknown | 5 (0.3) | 33 (1.6) | 26 (1.1) | 11 (0.5) | 75 (0.9) |
| Prior COVID-19 |  |  |  |  |  |
| No | 1565 (96.8) | 2031 (96.8) | 2375 (97.3) | 2054 (95.6) | 8025 (96.8) |
| Yes | 42 (2.6) | 42 (2.0) | 31 (1.3) | 64 (3.0) | 179 (2.2) |
| Unknown | 9 (0.6) | 26 (1.2) | 35 (1.4) | 30 (1.4) | 100 (1.2) |
| Household COVID-19 |  |  |  |  |  |
| No | 1573 (97.3) | 2014 (96.0) | 2276 (93.2) | 1983 (92.3) | 7846 (94.5) |
| Yes | 36 (2.2) | 49 (2.3) | 48 (2.0) | 137 (6.4) | 270 (3.3) |
| Unknown | 7 (0.4) | 36 (1.7) | 117 (4.8) | 28 (1.3) | 188 (2.3) |
| Close COVID-19 contact<sup>†</sup> |  |  |  |  |  |
| No | 1439 (89.0) | 1953 (93.0) | 1993 (81.6) | 1862 (86.7) | 7247 (87.3) |
| Yes | 30 (1.9) | 52 (2.5) | 62 (2.5) | 176 (8.2) | 320 (3.9) |
| Unknown | 147 (9.1) | 94 (4.5) | 386 (15.8) | 110 (5.1) | 737 (8.9) |
| COVID-19 vaccination<sup>†</sup> |  |  |  |  |  |
| No | 1089 (67.4) | 1633 (77.8) | 1925 (78.9) | 1904 (88.6) | 6551 (78.9) |
| Yes | 511 (31.6) | 452 (21.5) | 486 (19.9) | 236 (11.0) | 1685 (20.3) |
| Unknown | 16 (1.0) | 14 (0.7) | 30 (1.2) | 8 (0.4) | 68 (0.8) |
| Co-infections, positive/tested |  |  |  |  |  |
| HIV | 48/1586 (3.0) | 279/2075 (13.4) | 215/2362 (9.1) | 164/2008 (8.2) | 706/8031 (8.8) |
| Syphilis | 6/956 (0.6) | 11/538 (2.0) | 7/736 (1.0) | 22/1108 (2.0) | 46/3338 (1.4) |
| Hepatitis B | 0/400 (0.0) | 0/91 (0.0) | 0/41 (0.0) | 13/441 (2.9) | 13/973 (1.3) |
| Malaria | 190/1616 (11.8) | 18/512 (3.5) | 6/493 (1.2) | 27/464 (5.8) | 241/3085 (7.8) |
| COVID-19 risk behaviors<sup>§</sup> | n=988 | n=1382 | n=1690 | n=1334 | N=5394 |
| Mask use in public all or most of the time (self-report) | 428 (43.3) | 398 (28.8) | 755 (44.7) | 1090 (81.7) | 2671 (49.5) |
| Proper mask use during ANC visit | 574 (58.1) | 429 (31.0) | 873 (51.7) | 783 (58.7) | 2659 (49.3) |
| Travel outside district | 41 (4.1) | 47 (3.4) | 93 (5.5) | 100 (7.5) | 281 (5.2) |
| Travel outside Zambia | 6 (0.6) | 11 (0.8) | 5 (0.3) | 16 (1.2) | 38 (0.7) |
| Visit to church or mosque | 544 (55.1) | 993 (71.9) | 1201 (71.1) | 886 (66.4) | 3624 (67.2) |
| Visit to wedding or funeral | 236 (23.9) | 698 (50.5) | 381 (22.5) | 407 (30.5) | 1722 (31.9) |
| Visit to market or grocery store | 355 (35.9) | 1014 (73.4) | 1136 (67.2) | 1081 (81.0) | 3586 (66.5) |
| Indoor dining | 64 (6.5) | 437 (31.6) | 250 (14.8) | 352 (26.4) | 1103 (20.4) |
| Use of public transportation | 77 (7.8) | 918 (66.4) | 792 (46.9) | 989 (74.1) | 2776 (51.5) |
| ANC=antenatal care, IQR=interquartile range. <sup>†</sup>Close contact defined as being within 2 meters of someone outside the household suspected or confirmed to have COVID-19 for greater than 15 minutes; <sup>‡</sup>≥1 dose of any COVID-19 vaccine; <sup>§</sup>Responses restricted to February-September 2022 |  |  |  |  |  |

**Supplementary figure 1.**
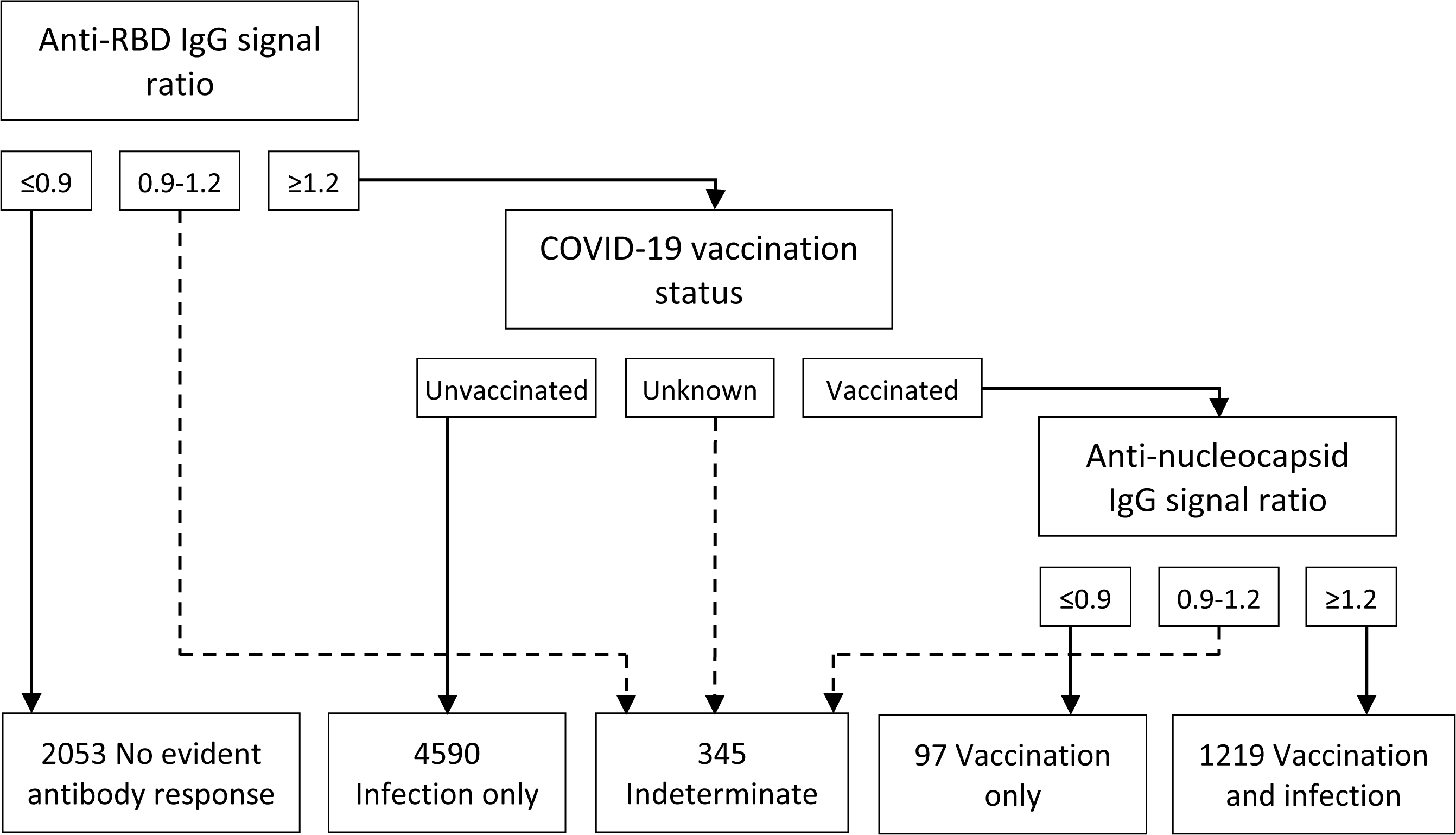
Antibody response categorization based on anti-RBD IgG, anti-nucleocapsid IgG, and COVID-19 vaccination status (N=8,304). The Tetracore FlexImmArray SARS-CoV-2 Human IgG Antibody Test used is a multiplex bead assay with three SARS-CoV-2 targets (i.e., nucleocapsid, receptor-binding domain (RBD) of spike, and fusion) to facilitate distinguishing between infection only, vaccination only, and combined vaccination and infection antibody responses when considered with vaccination history. Participants who reported receiving a Sinopharm or unknown vaccine were categorized as indeterminate due to the inability to distinguish between vaccination only and hybrid vaccination and infection antibody responses from inactivated or attenuated virus vaccines. Adapted from Duarte et al. 2021.

## References

1. Chipimo PJ, Barradas DT, Kayeyi N, et al. First 100 Persons with COVID-19 – Zambia, March 18–April 28, 2020. MMWR Morb Mortal Wkly Rep 2020; 69(42): 1547–48.

2. Zambia National Public Health Institute. Zambia COVID-19 Dashboard. Mar 14, 2023. https://www.arcgis.com/apps/dashboards/3b3a01c1d8444932ba075fb44b119b63 (accessed Apr 13, 2023).

3. Mulenga LB, Hines JH, Fwoloshi S, et al. Prevalence of SARS-CoV-2 in six districts in Zambia in July, 2020: a cross-sectional cluster sample survey. Lancet Glob Health 2021; 9: e773–81.

4. Shanaube K, Schaap A, Klinkenberg E, et al. SARS-CoV-2 seroprevalence and associated risk factors in periurban Zambia: a population-based study. Int J Infect Dis 2022; 118: 256–63.

5. National Institute for Communicable Diseases. SARS-CoV-2 seroprevalence in the Cape Town Metropolitan Subdistricts after the peak of infections. COVID-19-Special-Public-Health-Surveillance-Bulletin_Issue-5.pdf (accessed May 15, 2023).

6. Otieno NA, Azziz-Baumgartner E, Nyawanda BO, et al. SARS-CoV-2 infection among pregnant and postpartum women, Kenya, 2020-2021. Emerg Infect Dis 2021; 27(9): 2497–99.

7. Abdullahi A, Oladele D, Owusu M, et al. SARS-CoV-2 antibody responses to AZD1222 vaccination in West Africa. Nat Commun 2022; 13: 6131.

8. Sawry S, Le Roux J, Wolter N, et al. High prevalence of SARS-CoV-2 antibodies in pregnant women after the second wave of infections in the inner-city of Johannesburg, Gauteng Province, South Africa. Int J Infect Dis 2022; 125: 241–49.

9. Gonzalez R, Nhampossa T, Figueroa-Romero A, et al. SARS-CoV-2 seropositivity and HIV viral load among Mozambican pregnant women. J Acquir Immune Defic Syndr 2023; 92(2): 115–21.

10. Assefa N, Regassa LD, Teklemariam Z, et al. Seroprevalence of anti-SARS-CoV-2 antibodies in women attending antenatal care in eastern Ethiopia: a facility-based surveillance. BMJ Open 2021; 11: e055834.

11. Lucinde RK, Mugo D, Bottomley C, et al. Sero-surveillance for IgG to SARS-CoV-2 at antenatal care clinics in three Kenyan referral hospitals: Repeated cross-sectional surveys 2020-21. PLoS ONE 2022; 17(10): e0265478.

12. Duarte N, Yanes-Lane M, Arora RK, et al. Adapting serosurveys for the SARS-CoV-2 vaccine era. Open Forum Infect Dis 2021; 10.1093/ofid/ofab632.

13. UNICEF. COVID-19 Market Dashboard. Dec 21, 2020. https://www.unicef.org/supply/covid-19-market-dashboard (accessed Jun 12, 2023).

14. Lusaka Times. Zambia celebrates 770 percent COVID-19 vaccination achievement. Nov 2, 2022. https://www.lusakatimes.com/2022/11/02/zambia-celebrates-70-percent-covid-19-vaccination-achievement/ (accessed Jun 12, 2023).

15. Zambia Statistics Agency. Zambia demographic and health survey 2018. Jan 2020. https://www.zamstats.gov.zm/download/5400/?tmstv=1684233605&v=5436 (accessed Sep 25, 2020).

16. Makasa M, Fylkesnes K, Michelo C, Kayeyi N, Chirwa B, Sandoy I. Declining syphilis trends in concurrence with HIV declines among pregnant women in Zambia: Observations over 14 years of national surveillance. Sex Transm Dis 2012; 39(3): 173–81.

17. Gutman JR, Mwesigwa JN, Arnett K, et al. Using antenatal care as a platform for malaria surveillance data collection: study protocol. Malar J 2023; 22: 99.

18. Kangale C, Musunse M, Phiri-Chibawe C, et al. Routine malaria prevalence and intervention coverage estimates obtained by surveying antenatal care (ANC) attendees in Chadiza District, Eastern Province, Zambia: progress and lessons learned. ASTMH 2020. LB-5073

19. Tetracore Inc. Instructions for use: Tetracore FlexImmArray SARS-CoV-2 Human IgG Antibody Test. https://tetracore.com/wp-content/uploads/2021/04/FlexImmArraySARS-CoV-2IgGkitIFU_ver05122020_web.pdf (accessed Jan 9, 2023).

20. Tartof SY, Xie F, Yadav R, et al. Prior SARS-CoV-2 infection and COVID-19 vaccine effectiveness against outpatient illness during widespread circulation of SARS-CoV-2 omicron variant, US Flu VE Network. medRxiv 2023; published online Jan 11, 2023. 10.1101/2023.01.10.23284397 (preprint).

21. Mitchell KF, Carlson CM, Nace D, et al. Evaluation of a multiplex bead assay against single-target assays for detection of IgG antibodies to SARS-CoV-2. Microbiol Spectr; 10(3): 10.1128/spectrum.01054-22.

22. Zambia Statistics Agency. 2022 census of population and housing preliminary report. Dec 2022. https://www.zamstats.gov.zm/download/5578/?tmstv=1684233605&v=9742 (accessed Feb 14, 2023).

23. Lubeya MK, Kabwe JC, Mukosha M, et al. Maternal COVID-19 infection and associated factors: A cross-sectional study. PLoS ONE 2023; 18(3): e0281435.

24. Madhi SA, Kwatra G, Myers JE, et al. Population immunity and COVID-19 severity with Omicron variant in South Africa. N Engl J Med 2022; 386: 1314–26.

25. Madhi SA, Kwatra G, Myers JE, et al. Sustained low incidence of severe and fatal COVID-19 following widespread infection induced immunity after the Omicron (BA.1) dominant in Gauteng, South Africa: An observational study. Viruses 2023; 15(3): 597.

26. Altarawneh HN, Chemaitelly H, Ayoub HH, et al. Effects of previous infection and vaccination on symptomatic Omicron infections. N Engl J Med 2022; 387: 21–34.

27. Awandu SS, Ochieng AO, Onyango B, et al. High seroprevalence of Immunoglobulin G (IgG) and IgM antibodies to SARS-CoV-2 in asymptomatic and symptomatic individuals amidst vaccination roll-out in western Kenya. PLoS ONE 2022; 17(12): e0272751.

28. Konu YR, Conde S, Gbeasor-Komlanvi F, et al. SARS-CoV-2 antibody seroprevalence in Togo: a national cross-sectional household survey, May-June, 2021. BMC Public Health 2022; 22: 2294.

29. Arnaldo A, Mabunda N, Young PW, et al. Prevalence of severe acute respiratory syndrome coronavirus 2 (SARS-CoV-2) antibodies in the Mozambican population: a cross-sectional serologic study in 3 cities, July-August 2020. Clin Infect Dis 2022; 75(S2): S285–93.

30. Bergeri I, Whelan MG, Ware H, et al. Global SARS-CoV-2 seroprevalence from January 2020 to April 2022: A systematic review and meta-analysis of standardized population-based studies. PLoS Med 2022; 19(11): e1004107.

31. Yadouleton A, Sander A, Moreira-Soto A, et al. Limited specificity of serologic tests for SARS-CoV-2 antibody detection, Benin. Emerg Infect Dis 2021; 27(1): 233–37.

32. Chaponda D. Govt lifts wearing of masks in 22 districts. News Diggers! Aug 15, 2022. https://diggers.news/local/2022/08/15/govt-lifts-wearing-of-masks-in-22-districts/#:~:text=HEALTH%20Minister%20Sylvia%20Masebo%20has,%2C%20Chingola%2C%20Chifunabuli%20and%20Luanshya (accessed May 12, 2023).

33. Ripperger TJ, Urhlaub JL, Watanabe M, et al. Orthogonal SARS-CoV-2 serological assays enable surveillance of low-prevalence communities and reveal durable humoral immunity. Immunity 2020; 53: 925–33.

34. Alfego D, Sullivan A, Poirier B, Williams J, Adcock D, Letovsky S. A population-based analysis of the longevity of SARS-CoV-2 antibody seropositivity in the United States. EClinicalMedicine 2021; 36: 100902.

